# Synchronicity of P53 Mutation and Multiple High-Risk HPV Genotypes, and the Risk of Cervical Cancer among Women in Osogbo, Nigeria

**DOI:** 10.1101/2022.06.28.22277026

**Authors:** Musiliu A. Oyenike, Frederick O. Akinbo, Kamoru A. Adedokun, Hammed O. Ojokuku, Sikiru O. Imodoye, Lukman A. Yunus, Ismaila A. Lasisi, Abdulahi A. Jimoh

## Abstract

**Background:** In low-and middle-income countries, high burden of cervical cancer is associated with human papillomavirus (HPV) due to poor screening and diagnostic methods at early stage. Reports showed that there are discrepancies in data correlating HPV-infection with development of cervical cancer while the functional roles of *P53* oncogenic mutation are controversial. Furthermore, the molecular pathogenesis of multiple HPV-genotypes remains an open question. Thus, advancing investigations on HPV-associated cervical abnormalities would add to early diagnostic precision.

**Methods:** Two hundred (n=200) cervical samples were collected from apparently healthy, active adult women following an ethical approval. Laboratory analyses were conducted through cytological assessment and histochemistry screening using the Papanicolaou smear. PCR methods were used to characterize HPV-DNA genotypes and *P53* gene mutations. Positive cervical dysplasia cases were matched with HPV-DNA, and the HPV-genotypes were used to evaluate the prevalence of various HPV-subtypes and the risk of cervical cancer.

**Results:** Twenty-six (n=26) cervical dysplasia and fifty-one (n=51) HPV+ were identified comprising single and multiple genotypes. While nine cases (n=9) showed *p53* gene mutation with concurrent multiple high-risk HPV (hrHPV) genotypes, none of the single hrHPV genotypes had *p53* mutation. More so, individuals with coexisting *p53* mutation and multiple hrHPV-genotypes already manifesting cervical dysplasia were 22.2% of the group, while 77.8% had normal cervical architecture, the fate of whom was unknown during investigation. There was a higher cervical dysplasia among those with HPV oncogenes; there were connections between the HPV positivity with some genotypes (hrHPV16,18,31 and 33, respectively) and p53 mutation.

**Conclusion:** P53 gene mutation was independent of HPV-associated cervical abnormalities in single hrHPV-genotype, though other mechanistic drivers attributed to p53 dysfunction by E6 and E7 remain plausible. On the other hand, infection with multiple hrHPVs showed a concomitant predominance with the *P53* mutation, implying a potential interplay and an increased risk of cervical cancer.

## Background

According to a recent GLOBOCAN database, out of the infection-attributable cancer cases diagnosed worldwide, human papillomaviruses (HPV) caused about 31.1%, second to Helicobacter pylori (36.3%) [1]. HPV is the most common cause of cervical cancer, especially by high-risk HPV (hrHPV) genotypes such as HPVs 16, 18, 31, 33, 35, 39, 45, 51, 52, 56, 58, 59, 66, and 68, according to the World Health Organization (WHO); [2]. Also, hrHPVs are the most important risk factors present in 99.7% of the invasive cervical cancer, particularly among the young women [3]. In Nigeria, cervical cancer is one of the top five malignancies commonly diagnosed (excluding non-melanoma skin cancer). According to a recent global incidence analysis, cervical cancer accounts for 16.4% (equal to 12, 075 new cases) of the total 73, 417 new cancer cases among the female population, which include cancers of the breast, ovary, colorectum, non-Hodgkin lymphoma, and several others in Nigeria [4]. According to this report, Nigeria is one of the leading countries currently dealing with a high cervical cancer rate caused by HPV infection in females.

Although tremendous progress has been made in understanding HPV’s life cycle and role in the development of cervical cancer, the molecular events remain poorly understood and have been the subject of debate for mortality disparities around the world. A number of other factors, either directly or indirectly related to HPV infection, are associated with the development of cervical cancer in developing countries. These include a lack of knowledge, insufficient screening programs, inaccurate diagnosis, ineffective treatment methods, and increased exposure to risk factors. Generally, poor hygiene, smoking, oral contraceptive use, diethylstilbestrol (DES) exposure, and genetic predisposition have all been identified as common causes of cervical cancer in economically disadvantaged women [5]. From the knowledge of pathogenesis, cervical cancer develops after a long-term infection with the hrHPV, often after decades. Also, for cervical precancerous lesions to progress to invasive carcinoma, genetic and epigenetic alterations in host cell genes are involved.

Established knowledge shows that in many cancer cases a genetic predisposition is caused by mutations in tumor suppressor genes. P53 and retinoblastoma (Rb) are the two well-characterized tumor-suppressor genes, which encode proteins that repair damaged DNA. In cervical cancer patients, alterations in the Rb and p53 have been repeatedly implicated [6-8]. More importantly, P53 is known as the key player in the etiology of cervical cancer from a mechanistic standpoint. Unfortunately, several molecular interactions have been shown to suppress p53 activities, leading to cervical carcinogenesis [9]. P53 mutations, E6 and E7 malfunction, p53 degradation, overexpression of major p53 negative regulators like murine double minute 2 (MDM2) or MDM4, epigenetic dysregulation, and even alterations in TP53 mRNA splicing are just a few examples [6, 8]. Despite the fact that the tumor suppressor p53 is mutated in 50 percent of all human cancers [7] a simultaneous occurrence of p53 mutation and HPV infection, as well as a possible interaction that appears to be significantly related, remains doubtful. According to an earlier report, although p53 mutations are uncommon, HPV has been implicated in more than 90% of cervical malignancies [10]. To gain more insights, several studies further investigated the frequency of p53 mutations in cervix squamous cell carcinoma and adenocarcinoma, with varied outcomes. One of these studies examines the p53 mutation spectrum in cervical cancer cases from various geographical regions [11]. The findings show that the proportion of adenocarcinoma with mutated p53 ranged from 4% in North America to 19% in Asia, affecting six hot-spot codons of the gene, with three codons (175, 248 and 273) predominating in both types of the cancer [11].

A recent study of p53 gene mutation from African origin discovered various p53 gene mutations [12]. The study further explains that diverse environmental exposures and their vulnerabilities could be accountable for the divergence in polymorphism variants with connection to cervical oncogenesis. Other studies show that p53 is commonly inactivated by a variety of mechanisms, particularly in cancers with low p53 mutation rates, which could be a factor in HPV-positive cervical cancer, where p53 is ubiqutin-targeted for destruction by the E6 protein [13]. However, p53 mutation has been postulated as a possible unique pathway for oncogenesis in HPV-negative cervical cancer. More than one hrHPV genotype (multiple genotypes) are present in some individuals who are at a higher risk of cervical cancer, perhaps, causing synergistic effects and accelerating cervical architectural change, which may eventually leads to and hasten cervical cancer development.

Unfortunately, based on the studies above, the possible interaction of hrHPV genotypes with the concurrent p53 mutation could be unimaginable. Currently, the majority of available data focuses on either single hrHPV genotypes as a risk factor for cervical dysplasia or p53 mutation as a molecular driver in HPV-negative cervical cancer. More research is needed to better understand the molecular interplay of different hrHPV genotypes that orchestrate cervical abnormalities, especially in high-risk black populations. As a result, the goal of this study was to better understand the etiopathogenesis of cervical abnormalities in adult women in Osogbo metropolis, southwestern Nigeria, in relation to p53 mutation and hrHPV genotypes, in order to improve early diagnostic precision and reduce the risk of cervical cancer development.

## MATERIAS AND METHOD

### Study Area

This study was conducted at women attending the Obstetrics and Gynaecology clinics Ladoke Akintola University Teaching Hospital (LTH), Osogbo Central Hospital, and NasiruLahi-li-Fatihi Society of Nigeria (NASFAT), Health Center Osogbo, Osun State, southwestern Nigeria. Osogbo is a city in Nigeria with a population of 499,999 people and a land area of 2875 km^2^. It shares borders with Ikirun, Ilesa, Ede, Egbedore, Ibokun and Iragbiji [14]. The majority of the residents are Yoruba speakers and small-scale traders. As the capital of Osun State, Osogbo is the seat of government. Kwara State to the north, Oyo State to the west, Ogun State to the south, and Ondo and Ekiti States to the east, all share boundaries with Osun State.

### Study Population

A total of 200 cervical smears were collected from women in various parts of the city (as mentioned above). Participants ranged in age from 20 to 65 years old. Prior to the collection of the specimens, each participant was given a standardized questionnaire that asked about their bio-data and socio-demographic factors. Before collecting the specimen, each participant gave their written consent.

### Inclusion Criteria

All adult women between 20 years and above that consented to participate were recruited in this study.

### Exclusion Criteria

Participants that have other chronic diseases, those that refused consent and individuals less than 20 years were excluded from this study.

### Sample Size

The sample size (N) was determined using the formula N= Z^2^ P (1-p) / d^2^

Where; N = Minimum Sample Size

Z = confidence interval (1.96) from statistical table

P = prevalence rate (14.7%) [15].

d = desired level of significance taken as 0.05

N = Z^2^ P (1-p) / d^2^

N= (1.96)^2^x 0.147 (1-0.147) / 0.05^2^

N= 3.8416 × 0.147 × 0.853/ 0.0025

N= 3.8416 × 0.125391 / 0.0025

N= 0.4817020656 /0.0025

N= 192.68= 193 participants.

A minimum of 200 participants were recruited.

### Ethical Clearance

The protocol for this study was approved by the Osun State Health Research Ethics Committee (OSHREC) of the Ministry of Health, Osogbo, Osun State with reference number OSHREC/PRS/569T/158, 2019.

### Specimen Collection

Kavatkar et al. previously described a method for collecting cervical smears, which was used in this study [16]. Following sterile speculum dilatation of the vaginal wall, a cytobrush was introduced into the endocervical canal, turned clockwise, extracted, and placed in fixative. The mixture was allowed to sit for 30 minutes before being labeled [17]. It was then decanted and diluted with cellular solution after being placed into a spin tube and centrifuged at 1500rpm for 60 seconds. A 50ul portion of the diluted liquid was utilized to make a smear with a brush, while the rest was kept in the refrigerator for molecular analysis. The smear slides were air-dried for 3 to 5 minutes before being stained with Haematoxylin and Eosin. For DNA extractions, 500ul of refrigerated samples were placed in Eppendorf tubes, with the remaining samples being preserved in the refrigerator for future use [18].

### Processing of Specimen

#### Staining of Cervical Smear

Following the fixing, the Papanicoulaou staining technique was used to stain the smear. The smears were immersed in water for 4 minutes, stained with Harris haematoxylin, rinsed in water, differentiated in 0.5 % acid alcohol for 10 seconds, rinsed in water again, and blued in running tap water for 10 minutes. The stained smears were dipped in two changes of 90 % alcohol before staining in Orange G6 dye for 2 minutes. The stained smears were then rinsed three times with 95 % alcohol before being counterstained with Eosin Azure 50 solution for 4 minutes. The smears were rinsed in two changes of 95 % alcohol, dehydrated in two changes of absolute ethanol, cleared in xylene, and mounted in DPX [19]. The stained slides were reported according to the Bethesda System [20]. The reports were made as follow: normal without altered cells; atypical squamous cells of undetermined significance (ASC-US); atypical squamous cells – cannot exclude HSIL (ASC-H); low-grade squamous intraepithelial lesion (LSIL); and high-grade squamous intraepithelial lesion (HSIL). The cases with abnormal cytological diagnoses of HSIL and LSIL (Positives) were confirmed.

#### PCR Determination Method for HPV Gene

Following the manufacturer’s instructions, DNA was extracted using a Jena extraction kit. To ensure that enough DNA had been extracted, the extracted DNA was run on a 1% agarose gel. The samples were tested for the presence of HPV DNA using the GP5+/6+ primers MY09 (5′CGTCCM AARGGAWACTGATC-3′) and MY11 (5′-GCMCAG GGWCATAAYAATGG-3′), which target the conserved sequences in the HPV L1 region and may identify a variety of low and high risk mucosal HPVs. The samples were tested for the presence of HPV (the most commonly identified HR-HPVs in cervical cancer) using type specific primers [21].

### DNA Extraction

The full DNA extractions were carried out with the Jena extraction kit, following the manufacturer’s instructions with only minor modifications. The sample tubes were thawed, then vortexed and centrifuged for 1 minute each after adding 1000ul phosphate buffer. 300ul of lysate was pipetted into another tube, along with 2ul of proteinase K, vortexed for 15 seconds, and incubated for 1 hour at 55°C. 100ul protein precipitation was added, and the mixture was centrifuged for 1 minute. The supernatant was stirred 50 times and vortexed for 20 seconds after addition of 300ul of isopropanol. The supernatant was then discarded after centrifugation at 15,000 for 1 minute. 500ul of washing buffer was added to the tube, which was then inverted multiple times and air-dried at room temperature. Then, 50ul of hydrated solution was added for additional PCR amplication and other analyses [21, 22].

### Polymerase Chain Reactions for HPV and P53 Gene Mutation Analyses

In a 9600 thermocycler, polymerase chain reactions (PCRs) were performed. The PCR temperature cycles and duration for the reactions occurred as follow; denaturation at 94°C for 5 minutes, 94°C for 30 seconds, annealing at 56°C for 30 seconds, and extension at 72°C for 45 seconds. Each PCR reaction was preceded by a 5-minute denaturation step at 95°C, followed by a 5-minute extension step at 72°C. Depending on the sample volume reactions, the total number of PCR amplification cycles ranged from 30 to 40. 50ul final volume containing 50 mM KCl, 10 mM Tris-HCl pH 8.3, 200 M concentration of each dNTP, 1.5 mM MgCl2, 1ul of thermostable DNA polymerase (AmpliTaq DNA polymerase), and 15 pmol of each primer, was used. 2ul of the PCR product served as template for the nested PCRs. 10ul of the amplification products was analyzed by electrophoresis on 2% agarose gels and safragreen was used to stain [22].

### Detection of HPV-DNA Genotypes

To achieve precise HPV infection typing, type-specific nested multiplex Exon 6 primers were used. These primers were chosen from sequence alignments of the E6/E7 genes, with the goal of detecting sequence variations even among closely related genotypes. Nested primers were arranged in multiplex PCR primer cocktails to reduce the number of nested PCRs required to discriminate between a wide range of different HPV genotypes. The high-risk genotypes 16, 18, 31, 33, 35, 39, 45, 51, 52, 56, 58, 66, and 68 were identified using established primers. The primers were mixed into cocktails with four to five different primer pairs in each. The size of the nested PCR amplification product was determined by gel electrophoresis of molecular sizes 457, 322, 263, 398, 358, 280.151, 223, 229, 181, 274, 172, and 333 base pairs (Bp), respectively, to identify each HPV type present [22].

### Determination of HPV-DNA Genotyping Using PCR

For Taq amplification in the thermocycler 9600, a total reaction volume of 25 mL was prepared with 50 ng genomic DNA, 20 pmol of each primer, 200 lM of each dNTPs mixed with Taq buffer (10 mM, 384 Tris-HCl pH 8.3, 50 mM KCL), 3.0 mM MgCl2, and 3 units of Taq polymerase (New England, Biolabs). To guarantee complete extension of all PCR products, cycling settings were 3 minutes at 94°C for denaturation, 1 minute at 60 °C for annealing, and 7 minutes at 72 °C for final extension at 35 cycles. Gel electrophoresis was used to determine the PCR amplification product, which was done with a negative control and a 100 base pair molecular ladder.

### Detection of P53 Exon 6 Mutation

The P53 gene status, were evaluated after the DNA genotyping using the multiplex PCR method Mutations in the p53 gene occur most frequently within exons 5–8, which is the highly conserved DNA binding domain region [22]. Studies on p53 that included all exons have suggested that mutations outside exons 5–8 are rare in tumors [23]. The mutations of the p53 gene was verified by PCR-Single Strand Conformational Polymorphism (PCR-SSCP), which detects molecular changes in single-stranded DNA that cause changes in electrophoretic mobility through the following primers: Exon, 6 (5′-TGGTTGCCCAGGGTCCCCAG-3′) / (5′TGGAGGGCCACTGACAACCA–3′), 223 bp [22].

### Determination of p53 Gene Mutation

The p53 gene mutation was determined using 20 μl of DNA extracted lysates, a 9600 thermocycler, 10 mM Tris-HCl (pH 8.3), 2.0 mM MgCl2, 50 mM KCl, 0.1% Tween-20, 0.2 mM dNTP, 10 pM of each primer, and 2 units of AmpliTaq Gold polymerase (Perkin-Elmer). PCR reactions were denatured at 94°C for 30 seconds, annealed at 55°C for 30 seconds, and extended at 72°C for 30 seconds. Each PCR reaction was followed by a 7-minute extension at 72°C after a 5-minute denaturation at 95°C. Depending on the sample DNA, the total number of PCR amplification cycles ranged from 35 to 40. The size of the P53 gene amplification products size were determined using a gel electrophoresis equipment to identify electrophoresis bands. Specific forward and reverse primers selected for PCR reaction of p53 gene regulation are as follow:

p53

F = 50 -TGA AGT CTC ATG GAA GCC AGC-30

R = 50 -GCT CTTT TTC ACC CAT CTA CAG-30

## RESULTS

Out of the 200 women who took part in the study, 51 (25.5 percent) had HPV infection (Table 1). HPV-16, 18, 31, 33, 39, 66, and 68 were the HPV types discovered in this investigation, with HPV-31 having the highest frequency (Figs 1 - 3). In addition, the people aged 60 and up had the highest prevalence. Furthermore, participants who were traders had the highest risk of HPV infection, with the majority of them holding an elementary school certificate. Education and menopausal status, in particular, were found to have considerable connections with the risk of HPV infection. Religion affiliation, coital frequency, vaginal discharge, and menstruation pain, on the other hand, showed no link to HPV infection. Although the majority of the participants had normal cervical architecture, the proportion of HPV-infected people among those with abnormal cytology (dysplasia) was higher than in the normal group. Aside from that, multiple hrHPV-infected participants were seen. hrHPV 16 + 31 genotypes were the most predominant among them.

**Table 1:** Sociodemographic Characteristics of Study Participants.

| Relationship between age distribution and HPV infection |  |  |
| --- | --- | --- |
| Age | No. Infected/No Tested (%) | P-value |
| 20 – 29 | 5/27 (18.5) | 0.846 |
| 30 – 39 | 11/41(26.8) |  |
| 40 – 49 | 9/59(15.3) |  |
| 50 – 59 | 19/55(34.5) |  |
| 60 and above | 7/18(38.9) |  |
| Relationship between occupation and HPV |  |  |
| Occupation | No. Infected/No Tested (%) | P-value |
| Trader | 16/75 (21.3) | 0.417 |
| Student | 9/46 (19.6) |  |
| Civil servant | 19/54 (35.2) |  |
| House wife | 6/25 (24.0) |  |
| Association between educational status and HPV infection |  |  |
| Educational Status | No. Infected/No Tested (%) | P-value |
| None | 15/73 (20.5) | < 0.001 |
| Primary | 7/19 (36.8) |  |
| Secondary | 12/51 (23.5) |  |
| Tertiary | 17/57 (29.8) |  |
| Religion and HPV infection |  |  |
| Religion | No. Infected/No Tested (%) | P-value |
| Muslim | 31/129 (24.03) | 0.578 |
| Christians | 20/71 (28.2) |  |
| Relationship between menopausal status and HPV |  |  |
| Menopause | No. Infected/No Tested (%) | P-value |
| No | 29/142 (20.4) | 0.012 |
| Yes | 22/58 (37.9) |  |
| Relationship between coitus frequency and HPV |  |  |
| Coitus | No. Infected/No Tested (%) | P-value |
| None | 9/40 (22.5) | 0.619 |
| Weekly | 8/45 (17.8) |  |
| Monthly | 34/114 (29.8) |  |
| Association between abnormal vaginal discharge and HPV |  |  |
| discharge | No. Infected/No Tested (%) | P-value |
| No | 36/138 (26.1) | 0.249 |
| Yes | 15/62 (24.2) |  |
| Association between menstrual pain and HPV |  |  |
| menstrual pain | No. Infected/No Tested (%) | P-value |
| No | 43/166 (25.9) | 0.877 |
| Yes | 8/34 (23.5) |  |
| Association between dysplasia and HPV |  |  |
| Pap's smear | No. Infected/No Tested (%) | P-value |
| Normal | 41/174 (23.6) |  |
| Dysplasia | 10/26 (38.5) |  |
| Association between p53 mutation and HPV |  |  |
| p53 mutation | No of mutation per HPV status | P-value |
| Positive | 9/51 (HPV+) | <0.001 |
| Negative | 0/149 (HPV-) |  |
| Frequency of multiple hrHPV-genotypes among the participants |  |  |
| Combination of HPV types | Frequency (%) |  |
| 16 + 18 | 1(11.1%) |  |
| 18 + 31 | 2(22.2%) |  |
| 18 + 66 | 2(22.2%) |  |
| 16 + 31 | 4(44.4%) |  |

**Figure 1:**
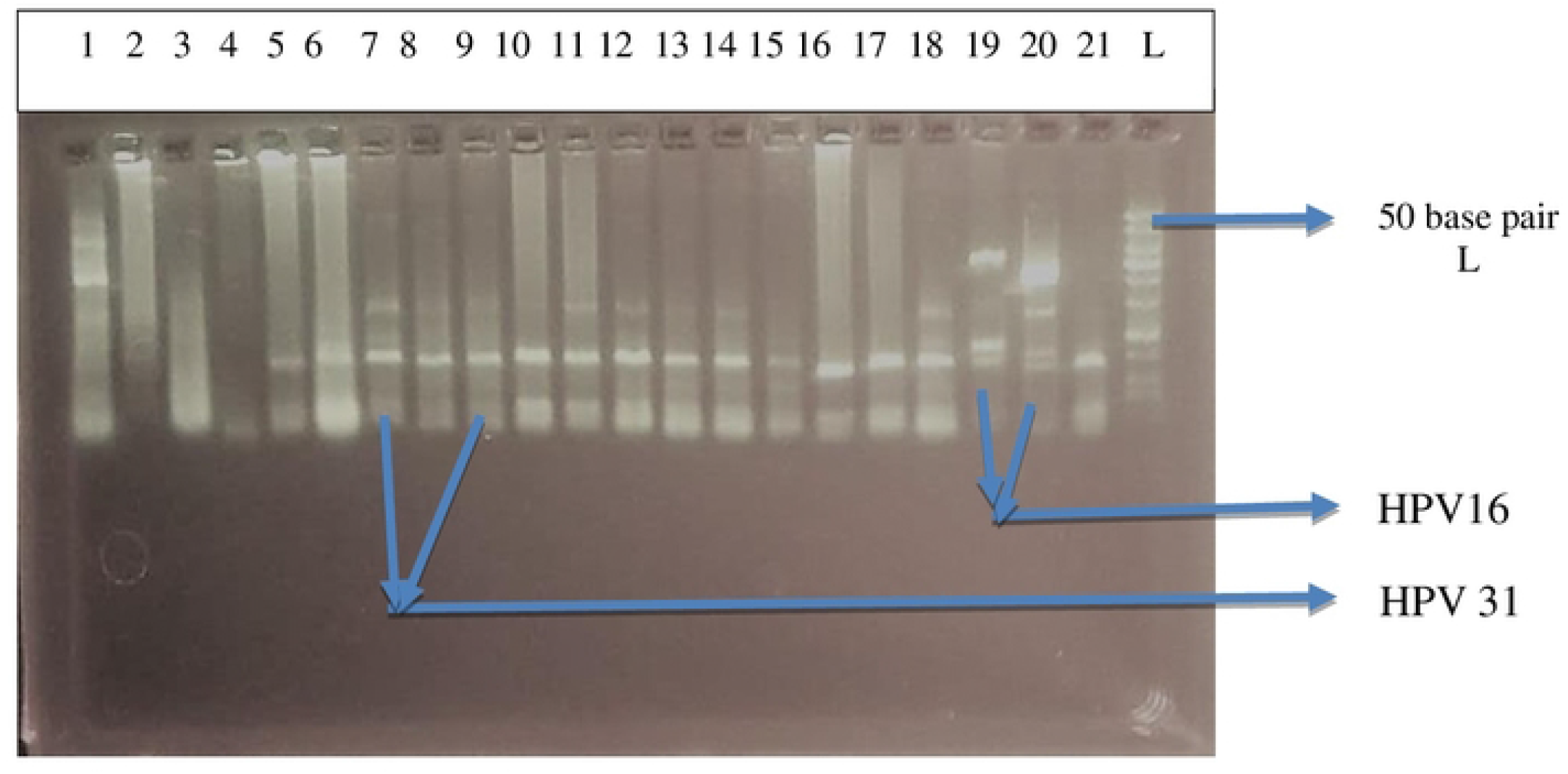
Image of agarose gel electrophoresis after multiplex PCR for HPV genotyping Lanes 7-14 and 18 = HPV 31 Lanes 1, 19 and 20 = HPV 16 Lanes 5, 6, 15 and 21 = HPV 18

**Figure 2:**
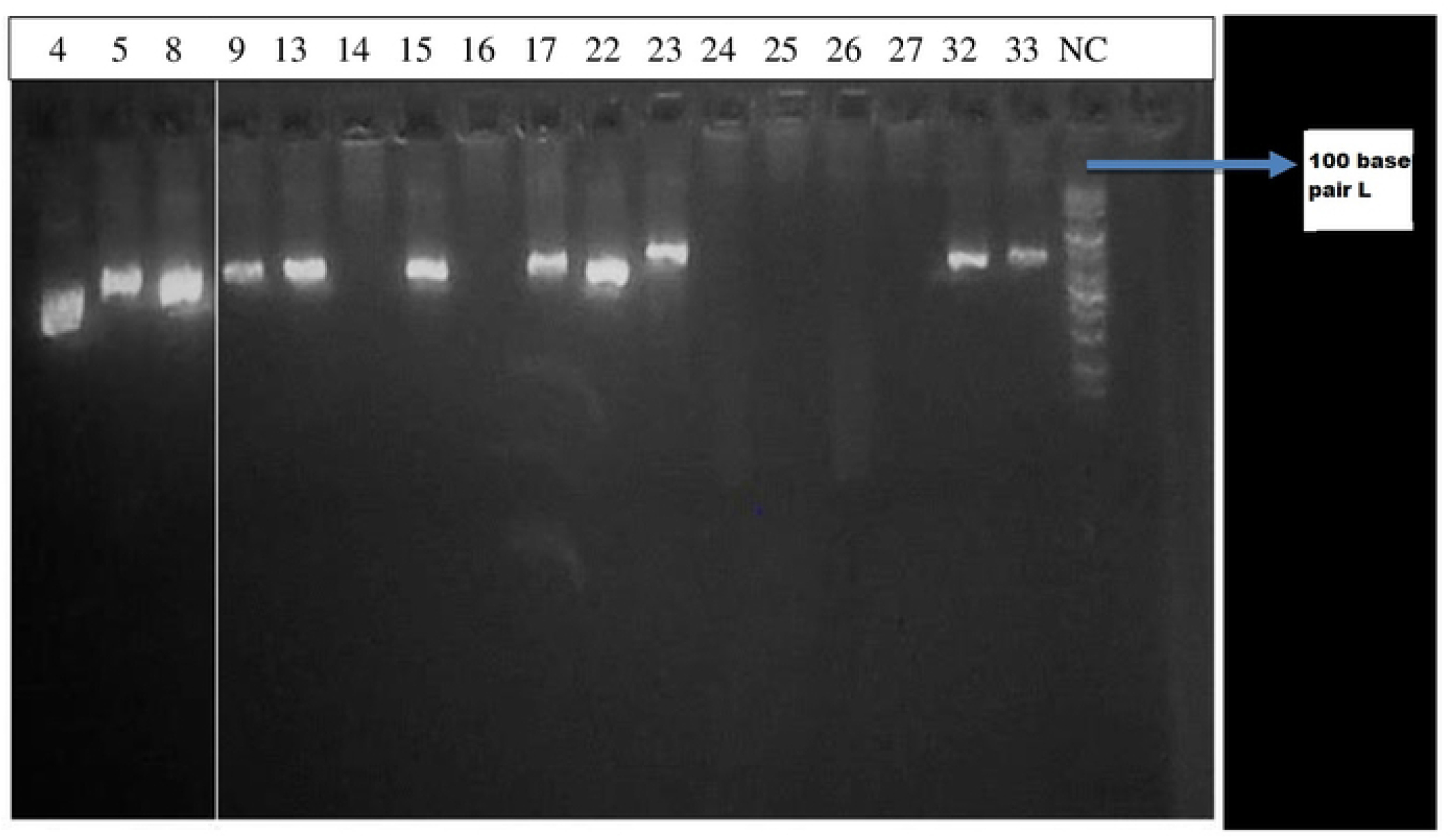
Image of agarose gel electrophoresis with primers targeting aprox 600 base pairs of HPV E6/7 genes Lanes 4,5,8,9,13,15,17,22,23,32 and 33 are positive for HPV DNA Lane L is 100 bp ladder Lane NC is the negative control

**Figure 3:**
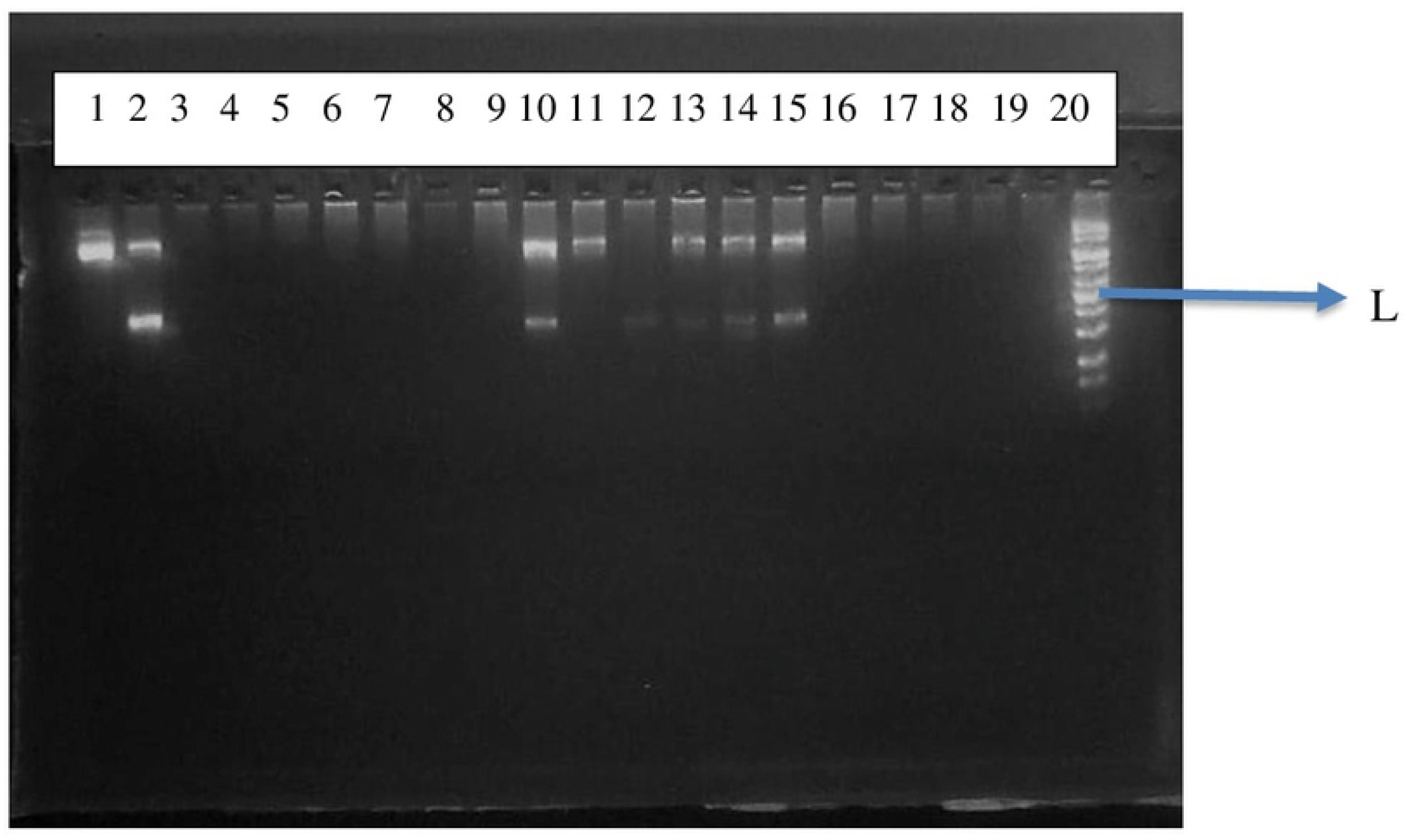
Image of Agarose gel electrophoresis after Exon 6 mutation (p53) detecting PCR Samples in lanes 1, 2, 10, 11, 12, 13, 14, and 15 all have exon 6 mutations

### Interpretation of result

Negative hrHPV genotype was 74.5 percent in this study, while single and multiple hrHPV genotypes were 21 percent and 4.5 percent, respectively. In addition, 10.7 percent of women had non-hrHPV-associated mild cervical dysplasia (cervical transformation via an HPV-independent pathway), while 11.9 percent had hrHPV-associated mild cervical dysplasia (precancerous changes associated with single hrHPV genotypes). Furthermore, the prevalence of hrHPV-associated severe cervical dysplasia (precancerous changes caused by a single hrHPV genotype) was 7.1 percent, while the prevalence of hrHPV-associated mild cervical dysplasia (precancerous changes due to multiple hrHPV genotypes) was 22.2 percent.The presence of a TP53 gene mutation was 100% positive for or in concordance with multiple hrHPV genotypes (both with cervical dysplasia and non-dysplasia). High-risk individuals with a combined p53 mutation and multiple hrHPV genotypes who had already manifested cervical dysplasia made up 22.2 percent of this group, while the remaining 77.8 percent had normal cervical tissue architecture, the fate of which was not known at the time of investigation (Table 2).

**Table 2:**
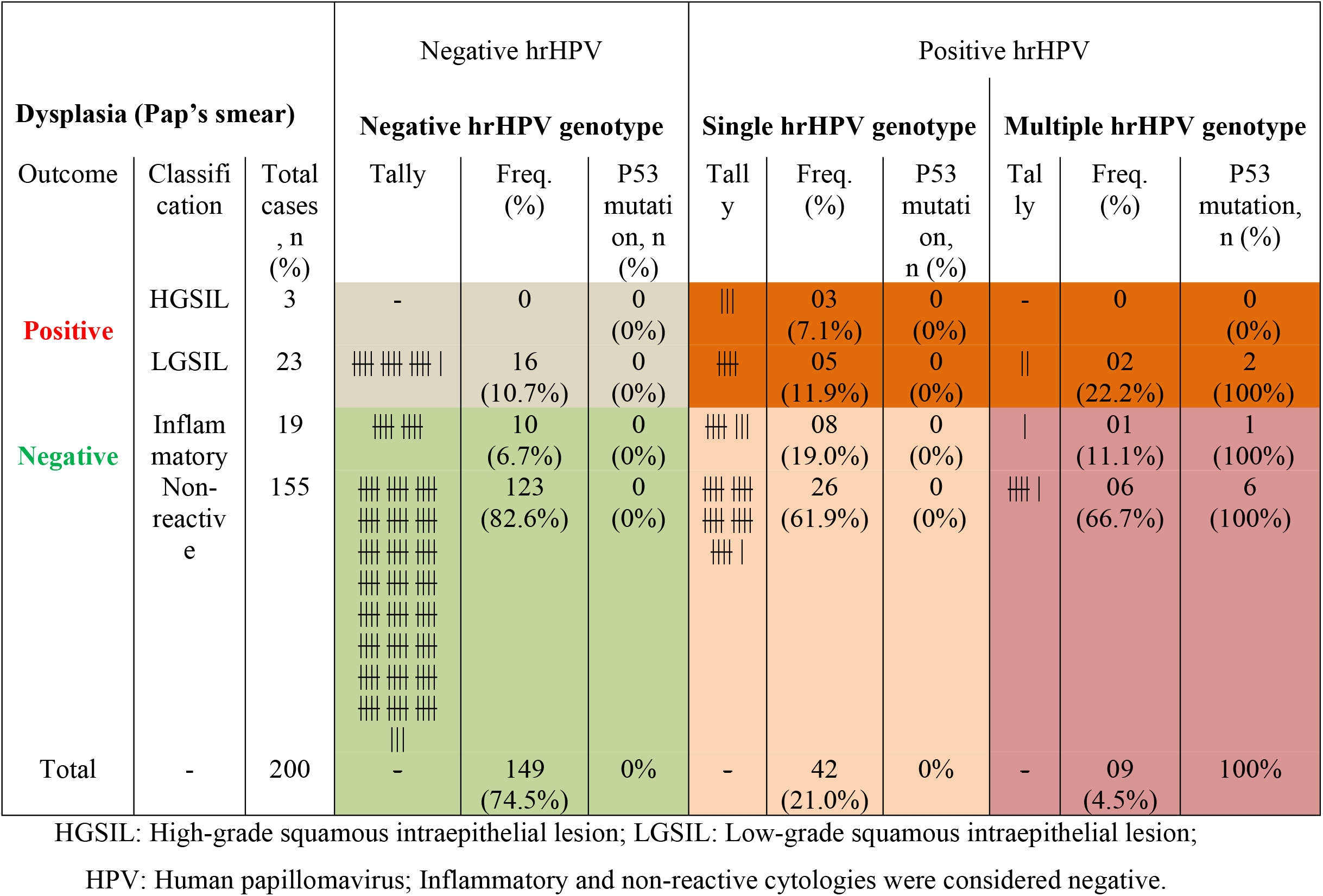
Molecular characterizations of hrHPV-genotypes and P53 mutation among the participants

## DISCUSSION

In this study, 200 cervical samples were investigated from consenting women in Osogbo metropolis, southwestern Nigeria, using molecular analyses and cervical cytology.We determined associations between high-risk human papillomavirus (hrHPV) infections and cervical dysplasia, as well as p53 mutation. We also investigated the relationship between these data and biological and socioeconomic factors.

The prevalence of cervical dysplasia (cervical abnormalities) was found to be 13% overall. This was in accordance with the study of Omole-Ohonsi [25] where 11.6% was reported in a five-year review of cervical cytology in Abakaliki, Nigeria. Investigating the relationship between cervical dysplasia and age, which is an established factor for cancer development, the current study found that people aged 60 and up had the highest frequency of hrHPV infection. Then came those aged 50 to 59, albeit there was no substantial difference when other age groups were taken into account, implying that other factors were at play. According to a report, the average age of cervical cancer diagnosis is 53 years, ranging from 44 to 68 years depending on the region [26]. Cervical cancer incidence increases with age. In other words, cervical cancer is uncommon between the ages of 20 and 24 years [27], which could explain why pre-cancerous (dysplasia) is more common as people become older, according to the current study. Pathologically speaking, cervical dysplasia are known as pre-cancerous cells that are yet to transform into cancerous cells, but which may have a risk of likelihood later in life, especially if not detected and treated early. It is worth noting that cervical cancer is more predominant among women groups who have poor access to screening [28]. Low practice of cervical cancer screening has been reported in Nigeria [29]. In the present study, it is suggestive that the abnormal positive cases for cytology might have age-related incidence who probably had not availed themselves for screening overtime. This is in agreement with a recent study, which suggests that older women from the poor-resource settings, and coincidentally from the rural areas, have increased risks to develop cervical cancer and are doubtful to have had screening [30]. This is further attributable to poor access to health services, ill-equipped health facilities and poor awareness [30].

Participants with lowest basic education level showed highest prevalence of HPV infections with HPV genotypes 16, 18, 31, and 33, respectively, while the least proportion was observed among those without formal education. This observation suggests that education significantly affected the prevalence of hrHPV infection among women in Osogbo metropolis. Previous report established that education has impacts on HPV infection *via* knowledge enrichment and creation of sexual right awareness [31]. As earlier mentioned, poor awareness has been repeatedly reported as an independent factor influencing HPV infection [30, 32]. Data from the literature suggest that educational interventions at high school levels could influence adolescents’ awareness, and offers both preventive knowledge and risk-taking behaviours from contracting HPV infection [32]. However, some sociodemographic variables, such as religion and occupation, had no association with the incidence of hrHPV infection, despite the fact that Muslim women were less infected compared to the Christian counterparts, and civil servants were slightly more infected. In terms of Islamic rites, vaginal cleansing may have a positive impact on HPV infection among the Muslim participants. Furthermore, a slightly higher proportion of participants had normal menstrual periods, despite having more hrHPV infections. Our findings suggest that having HPV-infection does not affect menstrual cycle. Unfortunately, the scope of our study did not take into account the menstrual phase. Although, the available information is still contradictory and inconsistent, according to one study, HPV detection is unrelated to menstrual cycle [33]. On the other hand, emerging evidence suggest that the menstrual cycle phase may influence the HPV-DNA detection [34]. The current study, however, highlights the need for additional research on the impact of hormones on papillomavirus genotype, pathogenicity, and epithelial tropism.

More so, the variations of hrHPV detection was cross-examined with the prevalence. The cumulative percentage of HPV infection was 25.5%, and the common oncogenic genotypes (often associated with genital cancers) were HPV16, HPV18, HPV31, HPV33, and HPV39, with HPV31 being the most predominant genotype. Various studies show inconsistent prevalence rates of HPV across the globe, and continental and local regions, using various methods of detection. In Ibadan, a neighbouring city in Nigeria, Neto et al. [35] recently reported 17.3% HPV prevalence using DNA consensus primer to identify fifteen different HPV genotypes, where only HPV-16, 18, 31, and 33 oncogenic subtypes are in accordance with the finding of the present study. All the same, the most predominant HPV genotype in the present study, HPV31, is in concordance with the outcome of Neto et al. [35]. Again, in Maiduguri, a Muslim dominated city in northern Nigeria, Kabir et al. [36] reported 69.8% prevalence rate of HPV using DNA detection method. Also, a robust review study of several low-resource countries by Nweke et al. [37] shows that sub-Saharan African has high prevalence rates of HPV infection predominated with three high-risk genotypes, HPV-16, 18, and 45, two of which were also detected in the present study. According to their age in relation with HPV prevalence rate, the oldest age range (60years and above) had highest prevalence of HPV infections followed by the second uppermost age range (50-59year). Report shows that older postmenopausal age women face challenges of persistent infection with HPV [38].

In relating sexual activities with the rate of contracting HPV, excluding other possible factors, this study further correlated coital frequency with HPV prevalence. Intriguingly, the women that had monthly regular coitus reported highest prevalence rate of 29.8%, while the weekly coitus had the least, 17.8% (Table 1). Of particular interest, the women with monthly coitus showed positivity for HPV31, HPV16, HPV18, HPV33 and HPV66 in that order of preponderance (with HPV 18 and 33 indicating equal rate and HPV66 the lowest). However, no statistical strength to show that regular sexual intercourse on monthly basis influence the disease in women who had the highest prevalence of HPV types 16, 18, 31 and 33 infections. Even though, on most occasions, HPV infection occurs through sexual intercourse [39, 40], albeit, HPV transmission is also possible by non-penetrative genital contacts and transmission *via* non-sexual routes such as from mother to baby during late prenatal and early post-natal periods, as well as through hand to genital contact [41] that might impact beyond coital frequency. It was therefore not surprising that the group with weekly coital frequency had a lower rate of hrHPV infection than monthly group, and that the group with sexual abstinence had higher hrHPV-infection rate than those with frequent intercourse. As a result, it was believed that other factors likely contributed to hrHPV infection in the current study and that it was unlikely that sexual activity had a significant impact.

A study of Chinese population in a rural area found oncogenic HPV types, especially for HPV16, 31, 33, 52, and 58, with HPV16 (19.1%) and HPV52 (16.5%) as the most preponderance and are often attributed to cervical intraepithelial neoplasia grade I and II (CINI&II) [42]. Apart from abnormal cytology report, a different but systematic review study of several reports from twenty three African countries reports that 4.4% and 2.8% of women with normal cytology have hrHPV16 and 18, respectively [43]. HPV16 and HPV18 are the most common HPVs detected worldwide, including many African countries, nonetheless, there are occasions where HPV-31 has been reported [35] similar with the present study.

Although HPV 16 has been widely reported as the most preponderant genotype, however, attributable to geographical variation, some studies report that HPV16 may not be the most common HPV genotype in some settings. Similarly, different HPV-genotypes have different clinical implications. Study shows that HPV 52 is the most common type in CIN II across all age categories [44], and has the highest preponderance as indicated in another study conducted in Zhejiang Province [45]. HPV 51, 52, and 31 are also more common in precancerous lesions than in invasive cervical cancer, despite the fact that HPV 16, 18, and 45 are more frequent in invasive cervical cancer than in any other grades of cervical malignancy [46].

Besides, many studies from Africa have observed multiple HPV infections [47]. However, those studies that identified multiple HPV types suggested the possibility of synergism and additivity, as well as an association with persistent infection and a higher risk of disease progression [48]. On the other hand, Salazar *et al*. [49] have shown that no significant difference occurs between single and multiple HPV infections. In this study, both HPV-associated and non-HPV associated cervical dysplasia (mild and severe precancerous changes) were observed (Table 2). Similarly, both single and multiple hrHPV genotypes were equally observed. Interestingly, all the cases with multiple hrHPV genotypes showed p53 inactivation with substantial portion manifesting low-grade squamous intraepithelial lesion (LGSIL). On the other hand, none among the participants with single HPV genotypes showed p53 dysfunction, albeit, some proportions already manifesting both high-grade squamous intraepithelial lesion (HGSIL) and LGSIL (Table 2). Meanwhile, dysfunction of p53 has been reported in several malignant tumours [50]. In particular, during cervical cancer progression, p53 inactivation occurs by degradation through the HPV oncoprotein E6 and E6-associated protein (E6AP) complex [51]. Kim et al. [52] demonstrated that TP53 gene mutations are infrequent in primary cervical cancer or precancerous lesions, and that HPV infection and TP53 gene mutation have no mutual relationship. On the other hand, our results of multiple hrHPV genotypes and P53 mutation occurring coincidentally, despite the lack of a discernible causal relationship, could indicate previously unreported molecular interplay in the etiology of cervical cancer - what we termed “synchronicity”.

Moreover, both hrHPV genotypes and P53 mutation are established risk factors for cervical cancer. Furthermore, despite the fact that the vast majority of this group (with many hrHPV genotypes) was still classed as normal cytology, there was a substantial number of aberrant cervical architecture (Table 2). Cervical cancer is a slow-growing malignancy. Because aberrant alterations in the cervix might take years or even decades to develop into invasive cancer cells, thus it is still unclear if the currently normal cytology will ultimately lead to a full-blown carcinoma.

Even though the majority of the subjects (74.5%) tested negative for the hrHPV genotype and without p53 mutation, a significant proportion of this group revealed a squamous intraepithelial lesion, non-HPV-associated dysplasia, the cause of which was unknown. This shows that cervical abnormalities were mediated by etiologic factors other than the p53 mutation and hrHPV-infection. In other words, there are other independent mechanisms exclusive of p53 inactivation, which might implicate development of cervical precancerous lesions as well as cervical cancer.

Literature shows that multiple genetic alterations accumulate in human cancers, which may involve inactivation of tumour suppressor genes [53] such as p53, oncogene activation, as well as loss of distinct chromosomal regions. However, base sequence mutation in p53 tumour suppressor gene occurs most frequently [54]. Albeit, in association with etiology of cervical cancer p53 gene mutations rarely occurs but only in 2-11% [52]. In the present study, p53 mutation was not reported among the participants either with single hrHPV genotypes or with negative for hrHPV infection; nonetheless, all participants with multiple HPV genotypes had p53 inactivation. On the contrary, this report also indicated that non-HPV-associated mild cervical dysplasia was still a striking figure, 10.7 % (table 2). In addition, both HGSIL and LGSIL do not necessary result in cancer if treatment is instituted aggressively. However, in the aftermath of cervical cancer development, our findings indicated that the idea that hrHPV causes about 99.7% of cancer as repeatedly reported [3, 55] or that p53 is exclusively involved in the molecular mechanism requires further investigation. As a result, the stakeholders should be aware of the possibility of a surge of millions of non-HPV-associated cervical cancer cases that might not benefit from vaccines, even though the current vaccination strategies are thought to eradicate cervical cancer in the near future.

## CONCLUSION

The p53 mutation and hrHPV infections were thought to be independent risk factors and determinants of cervical cancer development. Our findings, however, showed that the p53 mutation coexists in host tissue infected with multiple hrHPV genotypes. This suggests that multiple hrHPV infections and p53 susceptibility may interplay in a way that increases the risk of cervical cancer. However, in a single hrHPV genotype, p53 gene mutation was independent of HPV-associated cervical architectural abnormalities, though other mechanistic drivers attributed to p53 dysfunction by E6 and E7 remained plausible. Furthermore, non-HPV-associated cervical dysplasia was found in 10.7% of the participants in the current study while 99.7% of HPV-induced cervical cancer has been repeatedly reported in the literature. Our findings also implied that the PCR employing HPV DNA test is more sensitive than the Pap’s smear in detecting hrHPV at an early stage, even before cervical dysplasia develops. As a result, the HPV DNA test should be considered a baseline for early detection. Meanwhile, HPV infection is largely implicated in the pathogenesis of cervical cancer prompting the development of HPV vaccines. Therefore, putting these together, stakeholders should be aware of the possibility of an increase in the number of non-HPV-associated cervical cancer cases in less developed countries where efficient HPV screening methods are lacking.

## Limitation

The seeming number of participants who declined to take part was significant. The cultural and religious beliefs impacted the sample size significantly, considering the types of data required. The majority of the participants found it difficult to declare their sexual intercourse statuses, resulting in a large number of data gaps and thus excluded from the study. The reasons were considered to be in conjunction with unaccustomed use to declaring any sexual connexion openly or in any related ways, whether before marriage or out of the marital household in respect to Yoruba culture, and to some extent religiously.

## Data Availability

Data are available upon request.

## Acknowledgments

Prof. I. O. Ajayi of the University of Benin’s Department of Physiology deserves special thanks for his suggestions and advise on this project. We also acknowledge Dr. (Mrs) Faneye of the University of Ibadan, Department of Virology and Mrs. B. Farinloye of the University College Hospital, Pathology Department for their expertise and technical assistance during the analyses. Our gratitude goes to the staff members, from the Head of Department, Prof. (Mrs.) E.O. Osieme to other members most particularly Profs. M.A.Emokpae, I.N. Ibeh, F.E.Osaronsarje, H.B. Osadolor, M.A. Olungborva, E.O. Osime, E.B. Odigie for imparting this work positively.

## Author Contributions

Conceptualization: Frederick Akinbo, Musiliu Oyenike.

Data curation: Musiliu Oyenike.

Literature search: Musiliu Oyenike, Kamoru Adedokun

Formal analysis: Musiliu Oyenike.

Investigation: Musiliu Oyenike.

Methodology: Frederick Akinbo, Musiliu Oyenike, Kamoru Adedokun.

Statistical analyses: Hammed Ojokuku, Kamoru Adedokun.

Data interpretation: Kamoru Adedokun

Project administration: Frederick Akinbo, Musiliu Oyenike

Resources: Musiliu Oyenike.

Supervision: Frederick Akinbo.

Writing – original draft: Kamoru Adedokun.

Writing – review & editing: Kamoru Adedokun, Musiliu. Oyenike, Hammed Ojokuku, Sikiru Imodoye, Lukman Yunus, Ismaila Lasisi, and Abdulahi Jimoh.

